# Routine Testing for Chlamydia and Gonorrhea in an HIV Pre-Exposure Prophylaxis Program in Hanoi, Vietnam: Implications for Low- and Middle-Income Countries

**DOI:** 10.1101/2024.08.30.24312811

**Authors:** Paul C. Adamson, Hao T.M. Bui, Loc Q. Pham, Phuong T. Truong, Ngan T. Le, Giang M. Le, Jeffrey D. Klausner

## Abstract

**Objectives:** To assess the prevalence, anatomical distribution, and correlates of *Neisseria gonorrhoeae* (NG) and *Chlamydia trachomatis* (CT) infections within an HIV pre-exposure prophylaxis (PrEP) program in Hanoi, Vietnam.

**Design:** Cross-sectional, observational study

**Methods:** Between January-December 2022, HIV PrEP program clients who were male at birth, ≥16 years old, reported ≥1 male sex partner in the prior 12 months, were enrolled. A questionnaire collected sociodemographics, sexual behaviors, and clinical data. CT/NG testing was performed on self-collected urine, rectal, and pharyngeal specimens. Multivariate logistic regression was used to identify factors associated with infections.

**Results:** Among 529 participants, the prevalence of CT or NG was 28.9% (153/529). The prevalence of NG was 14.4% (76/529) and highest for pharyngeal infections (11.7%; 62/528), while for CT, the prevalence was 20.4% (108/529) and highest for rectal infections (15.0%; 74/493). Symptoms in the prior week were reported by 45.8% (70/153) of those with CT or NG infections. Among asymptomatic participants, there was a low prevalence of urethral CT (3.1%; 14/457) and NG (0.9%; 4/457). Condomless anal sex (aOR= 1.98; 95% CI: 1.27, 3.09) and sexualized drug use in the prior 6 months (aOR= 1.71; 95% CI: 1.09, 2.69) were associated with CT/NG infections.

**Conclusions:** The study found a high prevalence of CT/NG infections, particularly pharyngeal and rectal infections, within an HIV PrEP program in Hanoi, Vietnam. The findings suggest testing for urethral infections among asymptomatic individuals is of limited value. Further research is needed for STI prevention strategies and updated guidelines for CT/NG screening in HIV PrEP programs in low- and middle-income countries.

## Introduction

Sexually transmitted infections (STIs) are a significant public health challenge globally. *Neisseria gonorrhoeae* and *Chlamydia trachomatis* are the two most common bacterial STIs. Men who have sex with men (MSM) are disproportionately affected by STIs.[1,2] MSM on HIV pre-exposure prophylaxis (PrEP) have high frequencies of bacterial STIs. [3–5] Thus, major HIV PrEP guideline groups recommend routine screening for *C. trachomatis* and *N. gonorrhoeae* among MSM on HIV PrEP, including testing at multiple anatomic sites of infection.

In low- and middle-income countries (LMICs), HIV PrEP programs are expanding, yet data on STIs among MSM in HIV PrEP programs are limited. Understanding the epidemiology of STIs in these settings is important for planning and implementation of HIV PrEP programs. Moreover, in many LMIC settings, the availability of molecular *C. trachomatis* and *N. gonorrhoeae* testing is limited and the costs associated testing remain significant barriers for testing.[6,7] Identifying clinical and behavioral risk factors associated with those infections can help optimize diagnosis and prevention of STIs, particularly in LMIC settings where resources might be limited and there is ongoing rapid scale-up of HIV PrEP services.

Vietnam is a LMIC where HIV PrEP became available in 2018. Prior to that, baseline data from the Hanoi-MSM (HIM) study found a high prevalence of *C. trachomatis* (22%) and *N. gonorrhoeae* (12%) among MSM without HIV, who subsequently were enrolled into the pilot HIV PrEP project.[8] Currently, HIV PrEP guidelines in Vietnam recommend quarterly screening for *C. trachomatis* and *N. gonorrhoeae* at all sites with potential exposure[9]; however, the clients are responsible for covering the majority of the costs of those tests, which greatly limits testing uptake and available data on infections. The aims of this study are to fill gaps in research on STIs among MSM on HIV PrEP in Vietnam, since the rollout of HIV PrEP. The study objectives were to determine the prevalence of urethral, rectal, and pharyngeal *N. gonorrhoeae* and *C. trachomatis* infections within an HIV PrEP program in Hanoi, Vietnam, as well as to determine behavioral and clinical factors associated with *N. gonorrhoeae* and *C. trachomatis* infections.

## Methods

### Study design and population

This was an observational, cross-sectional study from January 2022 to December 2022. The study was conducted within the HIV PrEP program at the Sexual Health and Promotion (SHP) Clinic at Hanoi Medical University. Study participants were eligible if they were male sex at birth, aged 16 years or older, reported having sex with men or transgender women in prior 12 months, and were enrolled in, or presenting for enrollment into, the HIV PrEP program; exclusion criteria were if they had *C. trachomatis* or *N. gonorrhoeae* testing done in the prior 3 months, unless they reported acute STI symptoms on the day of enrollment. The exclusion criteria were included to align closely with the HIV PrEP program recommendations. Participants were allowed to participate in the study more than once. Eligible participants were approached in the clinic and the study’s objectives and procedures were explained by study research staff.

### Data collection

Demographic, behavioral, and clinical characteristics were collected using a tablet self-administered survey. Sexual behaviors were self-reported and included: number of sex partners in the prior month as the combined number of male, female, and transgender sex partners, anal sex position(s), condomless anal intercourse in the prior month, group sex (defined as having more than one sex partner at the same encounter), or having sex with partners met via mobile apps in the prior 6 months. Sexualized drug use was considered if the participant reported using a substance (heroine, ketamine, ecstasy, methamphetamine, gamma hydroxybutyrate or gamma butyrolactone, poppers, or prostaglandin E medications) in the prior 6 months to enhance sexual pleasure. Poppers are commonly included in the definition of sexualized drug use in the Asia Pacific region.[10] History of STIs in the prior three months was obtained by self-report.

Participants were asked about any genitourinary, rectal, or pharyngeal symptoms in the prior one week. Genitourinary symptoms were classified as any of the following: pain with urination, discharge, bleeding, pruritis, testicular pain, lymphadenopathy, or ulcers. Rectal symptoms were classified as any of the following: tenesmus, dyschezia, pruritis, bleeding, discharge, ulcers, or diarrhea. Pharyngeal symptoms included pain or itching in the throat.

### Sample collection, testing, and treatment

Study participants received instructions on providing rectal, pharyngeal, and urine specimens for testing by study staff and with visual aids. Clinician-collected samples were obtained if participants preferred or were unable to perform self-collection. Study specimens were collected using either the Alinity m multi-Collect Specimen Collection Kits (Abbott Molecular, USA) or cobas PCR Urine or Swab Sample Kits (Roche Diagnostics, Branchburg, NJ, USA).

Specimens were stored in the clinic and transported daily to the laboratory. Testing was performed using either the Alinity m STI Assay (Abbott Molecular, USA) or the cobas 4800 CT/NG assay (Roche Diagnostics, Branchburg, NJ, USA) according to manufacturer’s instructions. Test results for *C. trachomatis* and *N. gonorrhoeae* were either positive, negative, or inconclusive, in the event of failed internal controls or the presence of inhibitors. Participants were informed of their test results. For those found to have infections, free treatment was offered. For *C. trachomatis* infections, doxycycline 100mg by mouth twice daily for 7 days was provided. For *N. gonorrhoeae* infections, ceftriaxone 500mg intramuscular injection once was the preferred treatment offered; cefixime 800mg by mouth once was provided as an alternative treatment.

### Data analysis

The primary outcome was the prevalence of *C. trachomatis* or *N. gonorrhoeae* infections at different anatomic sites. The study sample size was determined based on available resources, staffing, and a defined study duration (2022). Rather than relying on formal sample size calculations, recruitment was intentionally inclusive of all eligible study participants, that could be enrolled based on research timing and staffing. A sample size of 500 would have a reasonable estimate of infection prevalence with a margin of error <5%. Descriptive statistics were performed for demographic, behavioral and clinical data; percentages for categorical variables, or median and interquartile range for continuous variables, were reported. Categorical variables between those with and without *C. trachomatis* or *N. gonorrhoeae* infections were compared using Pearson’s Chi-squared test and Fisher’s exact test, when frequencies were low, and nonparametric continuous variables were compared using Wilcoxon rank-sum tests.

Logistic regression modeling was used to evaluate factors associated with *N. gonorrhoeae* and *C. trachomatis* infections separately as well as the outcome of having either infection. Variables with p-values < 0.20 in univariate comparisons were included in the multivariable logistic regression models; except for age, which was included in the final models. The number of sex partners in the prior month was dichotomized to above or below the median (0-1 and ≥2) to improve interpretation and model fit. For participants with multiple study visits, data analysis was restricted to their first participation. A variance inflation factor was used to measure for multicollinearity between variables in the multivariate logistic regression. Variables with missing data were excluded from the logistic regression models. P-values ≤ 0.05 was defined as statistical significance. All data analyses were performed using STATA version 18 (StataCorp LLC, College Station, TX, USA).

### Ethics

The study was approved by the Institutional Review Boards of Hanoi Medical University (HMUIRB580), the University of California, Los Angeles, and the University of Southern California. All study participants provided written informed consent. People aged 16 years or older are eligible for the HIV PrEP program, thus study eligibility included these individuals, who could participate in the study with assent and parental consent. Participants were compensated 200,000 VND (∼9 USD) for their time and effort of study participation.

## Results

### Participant Characteristics

From January to December 2022, there were 529 participants enrolled into the study among 775 approached and 538 screened. The primary reasons for refusal of those who were approached, but who did not undergo screening, was “too busy/not enough time” or “will consider next time.” Multiple visits occurred for 134 participants: 116 with two, 16 with three, and two with four visits. Among the study participants, 81.6% (432/529) identified as a man, 1.7% (9/529) identified as transgender woman, and 16.6% (88/529) identified as gender non-conforming. (Table 1). The median age among all participants was 25.1 years (IQR: 21.7-29.5). There were 29.1% (154/529) currently undergoing university or post-secondary training, while 61.1% (323/529) had completed university or post-secondary training.

**Table 1.** Baseline demographic, behavioral, and clinical characteristics of 529 participants enrolled from an HIV PrEP program clinic in Hanoi, Vietnam.

|  | Overall<br>(N = 529) | Positive for *C. trachomatis* (N=108) |  | Positive for *N. gonorrhoeae* (N=76) |  | Positive for *C. trachomatis* or *N. gonorrhoeae* (N=153) |  |
| --- | --- | --- | --- | --- | --- | --- | --- |
|  | n (%) | n (%) | p-value | n (%) | p-value | n (%) | p-value |
| **Median Age (IQR)** | 25.1 (21.7-29.5) | 25.3 (21.1 – 29.7) | 0.869<sup>†</sup> | 24.8 (22.1 – 27.2) | 0.281<sup>†</sup> | 25.1 (21.5 – 28.3) | 0.336<sup>†</sup> |
| **Age group** |  |  | 0.644 |  | 0.243 |  | 0.834 |
| 16- 24 years old | 261 (49.3) | 51 (19.5) |  | 38 (14.6) |  | 75 (28.7) |  |
| 25- 34 years old | 221 (41.8) | 45 (20.4) |  | 35 (15.8) |  | 66 (29.9) |  |
| 35 - 63 years old | 47 (8.9) | 12 (25.5) |  | 3 (6.4) |  | 12 (25.5) |  |
| **Gender self-identification** |  |  | 0.126<sup>‡</sup> |  | 0.316<sup>‡</sup> |  | 0.232<sup>‡</sup> |
| Man | 432 (81.7) | 85 (19.6) |  | 58 (13.4) |  | 121 (28.0) |  |
| Transgender woman | 9 (1.7) | 0 |  | 1 (11.1) |  | 1 (11.1) |  |
| Other/Unsure* | 88 (16.6) | 23 (26.1) |  | 17 (19.3) |  | 31 (35.2) |  |
| **Education** |  |  | 0.687 |  | 0.942 |  | 0.797 |
| High school or below | 52 (9.8) | 13 (25.0) |  | 8 (15.4) |  | 17 (32.7) |  |
| In post-secondary training | 154 (29.1) | 31 (20.1) |  | 21 (13.6) |  | 45 (29.2) |  |
| Completed post-secondary training or higher | 323 (61.1) | 64 (19.8) |  | 47 (14.6) |  | 91 (28.2) |  |
| **Monthly income in million VND (median, IQR)** | 9.0 (5 – 15) | 8.5 (5-14) | 0.813<sup>†</sup> | 8 (4.5 – 14.5) | 0.566<sup>†</sup> | 8 (5 – 14.5) | 0.493<sup>†</sup> |
| **Employed** | 419 (79.2) | 84 (20.0) | 0.682 | 63 (15.0) | 0.392 | 123 (29.4) | 0.668 |
| **Gender of sex partners prior 6 months** |  |  | 0.711<sup>‡</sup> |  | 0.327<sup>‡</sup> |  | 0.502<sup>‡</sup> |
| Men only | 474 (89.6) | 96 (20.3) |  | 73 (15.4) |  | 141 (29.7) |  |
| Women only | 13(2.5) | 2 (15.4) |  | 1 (7.7) |  | 2 (15.4) |  |
| Men and women | 33 (6.2) | 9 (27.3) |  | 2 (6.0) |  | 9 (27.3) |  |
| No sex partners | 9 (1.7) | 1 (11.1) |  | 0 |  | 1 (11.1) |  |
| **Number of sex partners in prior 1 month (n = 520)<sup>#</sup>** | 1 (1 – 2) | 2 (1 – 3) | <0.001<sup>†</sup> | 2 (1 – 2) | 0.150<sup>†</sup> | 2 (1 – 3) | <0.001<sup>†</sup> |
| **Anal sex with male partners in prior 1 month (n = 447)<sup>#</sup>** | 396 (88.4) | 95 (24.0) | 0.043 | 64 (16.2) | 0.219 | 131 (33.1) | 0.043 |
| **Anal sex position with male partner (n = 396)<sup>#</sup>** |  |  | 0.633 |  | 0.109 |  | 0.385 |
| Insertive anal sex always | 137 (34.6) | 29 (21.2) |  | 22 (16.1) |  | 44 (32.1) |  |
| Receptive anal sex always | 149 (37.6) | 38 (25.5) |  | 18 (12.1) |  | 45 (30.2) |  |
| Both insertive and receptive | 110 (27.8) | 28 (25.5) |  | 24 (21.8) |  | 42 (38.2) |  |
| **Condomless anal sex in prior 1 month (n = 396)<sup>#</sup>** | 203 (51.3) | 63 (31.0) | 0.001 | 39 (19.2) | 0.091 | 83 (40.9) | 0.001 |
| **Vaginal sex with female partners in prior month (n = 39)<sup>#</sup>** | 38 (7.2) | 8 (21.1) | 0.064 | 2 (5.3) | 0.814 | 8 (21.1) | 0.064 |
| **Deep kissing in prior week (n = 302)<sup>#</sup>** | 260 (86.1) | 61 (23.5) | 0.700 | 41 (15.8) | 0.518 | 85 (32.7) | 0.823 |
| **Any group sex, prior 6 months** | 65 (12.3) | 13 (20.0) | 0.929 | 14 (21.5) | 0.078 | 22 (33.8) | 0.350 |
| **Sexualized drug use with** | 235 (44.4) | 67 (28.5) | <0.001 | 40 (17.0) | 0.120 | 87 (37.0) | <0.001 |

|  | Overall<br>(N = 529) | Positive for *C. trachomatis* (N=108) |  | Positive for *N. gonorrhoeae* (N=76) |  | Positive for *C. trachomatis*<br>or *N. gonorrhoeae*<br>(N=153) |  |
| --- | --- | --- | --- | --- | --- | --- | --- |
|  | n (%) | n (%) | p-value | n (%) | p-value | n (%) | p-value |
| **poppers or erectile dysfunction medications, prior 6 months** |  |  |  |  |  |  |  |
| **Any sexualized drug use, prior 6 months** | 239 (45.4) | 68 (28.4) | <0.001 | 40 (16.7) | 0.169 | 88 (36.8) | <0.001 |
| **Met sex partners via mobile apps, prior 6 months** | 297 (56.1) | 66 (22.2) | 0.244 | 55 (18.5) | 0.002 | 97 (32.7) | 0.032 |
| **Pharyngeal symptoms in prior week** | 126 (23.8) | 21 (16.7) | 0.232 | 20 (15.9) | 0.581 | 33 (26.2) | 0.438 |
| **Rectal symptoms in prior week** | 87 (16.5) | 25 (28.7) | 0.035 | 15 (17.2) | 0.403 | 29 (33.3) | 0.321 |
| **Genitourinary symptoms in prior week** | 72 (13.6) | 17 (23.6) | 0.469 | 15 (20.8) | 0.092 | 26 (36.1) | 0.148 |
| **Any genitourinary symptoms at the day of enrollment** | 14 (2.7) | 7 (50.0) | 0.012<sup>‡</sup> | 8 (57.1) | <0.001<sup>‡</sup> | 12 (85.7) | <0.001<sup>‡</sup> |
| **Any symptoms in prior week** | 217 (41.0) | 49 (22.6) | 0.303 | 41 (18.9) | 0.013 | 70 (32.3) | 0.158 |
| **Self-reported STI diagnosis in the prior 3 months** | 29 (5.5) | 6 (20.7) | 0.970 | 6 (20.7) | 0.318 | 9 (31.0) | 0.796 |
| **Used antiseptic mouthwash in the previous month** | 286 (54.1) | 65 (22.7) | 0.152 | 38 (13.3) | 0.442 | 84 (29.4) | 0.805 |

Note: Column percentages shown for first column. Remaining values are shown as row percentages.
\*Participants who were gender nonconforming or gender incongruent, or unsure of their gender identity.
<sup>†</sup>Wilcoxon rank-sum (Mann-Whitney) test was performed
<sup>‡</sup>Fisher's exact test was performed, Chi-Squared tests were performed otherwise
<sup>§</sup>Variables that contains missing values because "Don't know" or "Don't remember" choices were treated as missing.
<sup>#</sup>Sample size was lower than 529 due to skip pattern (e.g., sample size for "Condomless anal sex" was among people who reported engaging in anal sex)

In the prior six months, 95.8% (507/529) reported only male sex partners; the median number of sex partners in the prior month was 1 (IQR: 1-2). (Table 1). Anal sex in the prior 6 months was reported by 88.4% (396/529), of which 51.2% reported condomless anal sex in the prior month. In the prior 6 months, group sex was reported by 12.3% (65/529) and sexualized substance use was reported by 45.4% (240/529). A history of a STI in the prior 3 months was reported by 5.5% (29/529), with 69.0% (20/29) of those being syphilis. Additional sexual behavior data are shown in Table 1.

### C. trachomatis and N. gonorrhoeae infections

All participants submitted a urine sample for testing, a pharyngeal specimen was not collected for one (0.2%) participant, while 11 (2.1%) participants refused rectal specimen collection. In total, 28.9 % (153/529) participants had *C. trachomatis* or *N. gonorrhoeae* infections at any anatomical site. The prevalence of C. *trachomatis* was 20.4% (108/529) and 14.3% (76/529) for *N. gonorrhoeae*; 5.9% (31/529) of participants had *C. trachomatis* and *N. gonorrhoeae* co-infections (Table 1). Among the 153 participants with either *C. trachomatis* or *N. gonorrhoeae* infections, the median age was 25.1 years (IQR: 21.5 – 28.3). The median number of sex partners in the prior month was 2 (IQR: 1-3), 14.2% (22/155) reported group sex in the prior month, and 60.1% (94/155) reported sexualized substance use. Of the 153 with *C. trachomatis* or *N. gonorrhoeae* infections, 45.8% (70/153) reported any symptoms in the prior week; 33 (21.6%) reported pharyngeal symptoms, 29 (19.0%) reported rectal symptoms, and 26 (17.0%) reported urethral symptoms. More participants with *C. trachomatis* or *N. gonorrhoeae* infections reported having symptoms on the day of enrollment, compared to those without infections (9.2% vs. 3.0%, p=0.001).

*C. trachomatis* and *N. gonorrhoeae* infections by anatomic site are shown in Table 2. The prevalence of *C. trachomatis* was highest in the rectum (15.0%; 74/493) and 3.4% (18/529) had infections in multiple anatomic sites. For *N. gonorrhoeae*, pharyngeal site infections were most prevalent (11.7%; 62/528) and 6.0% (32/529) had infections at multiple anatomic sites. Among participants with either *C. trachomatis* or *N. gonorrhoeae*, the prevalence of rectal infections was 19.1% (94/493) and 9.3% (49/529) had infections at multiple anatomic sites. For rectal specimens, 24 (4.5%) were inconclusive for both infections and one additional result was inconclusive for *C. trachomatis*.

**Table 2.** Prevalence of *Chlamydia trachomatis* and *Neisseria gonorrhoeae* infections by anatomic site among 529 participants enrolled from an HIV PrEP program clinic in Hanoi, Vietnam.

| Anatomic Site | *C. trachomatis*<br>(n/N*, %) | *N. gonorrhoeae*<br>(n/N*, %) | *C. trachomatis* or<br>*N. gonorrhoeae*<br>(n/N*, %) | *C. trachomatis* and<br>*N. gonorrhoeae*<br>(n/N*, %) |
| --- | --- | --- | --- | --- |
| **Total Infections (Any Site)** | 108 (20.4) | 76 (14.4) | 153 (28.9) | 31 (5.9) |
| **Single Site Infections** |  |  |  |  |
| Pharyngeal only | 19/528 (3.6) | 32/528 (6.1) | 41/528 (7.7) | 3/528 (0.6) |
| Rectal only | 57/493 (11.6) | 10/494 (2.0) | 52/493 (10.6) | 4/493 (0.8) |
| Urethral only | 14/529 (2.7) | 2/529 (0.4) | 11/529 (2.1) | 0/529 (0) |
| **Multisite Infections** |  |  |  |  |
| Pharyngeal and rectal | 9/493 (1.8) | 18/494 (3.6) | 26/494 (5.3) | 13/493 (2.6) |
| Pharyngeal and urethral | 1/528 (0.2) | 5/528 (1.0) | 7/528 (1.3) | 3/528 (0.6) |
| Urethral and rectal | 7/493 (1.4) | 2/494 (0.4) | 7/493 (1.4) | 1/493 (0.2) |
| Pharyngeal, rectal, and urethral | 1/493 (0.2) | 7/494 (1.4) | 9/494 (1.8) | 7/493 (1.4) |
| **Any pharyngeal** | 30/528 (5.7) | 62/528 (11.7) | 83/528 (15.7) | 26/528 (4.9) |
| **Any rectal** | 74/493 (15.0) | 31/493 (6.3) | 94/493 (19.1) | 25/493 (5.1) |
| **Any urethral** | 23/529 (4.3) | 16/529 (3.0) | 34/529 (6.4) | 11/529 (2.1) |

The prevalence of *C. trachomatis* or *N. gonorrhoeae* infections by anatomic site, stratified by reported symptoms in the prior week are shown in Figure 1. Among those reporting rectal symptoms in the prior week, the prevalence of having a rectal infection with *C. trachomatis* or *N. gonorrhoeae* was 27.2% (22/81), followed by 25.0% (18/72) for urethral symptoms and urethral infections, and 18.3% (23/126) for pharyngeal symptoms and pharyngeal infections. Among those who did not report symptoms in the prior week, the prevalence of urethral *C. trachomatis* was 3.1% (14/457) and 0.9% (4/457) for *N. gonorrhoeae*. For both infections, the prevalence of both rectal and urethral infections were higher among those reporting symptoms in the prior week; there were no differences in the prevalence of pharyngeal infections by symptom status for either infection.

**Figure 1.**
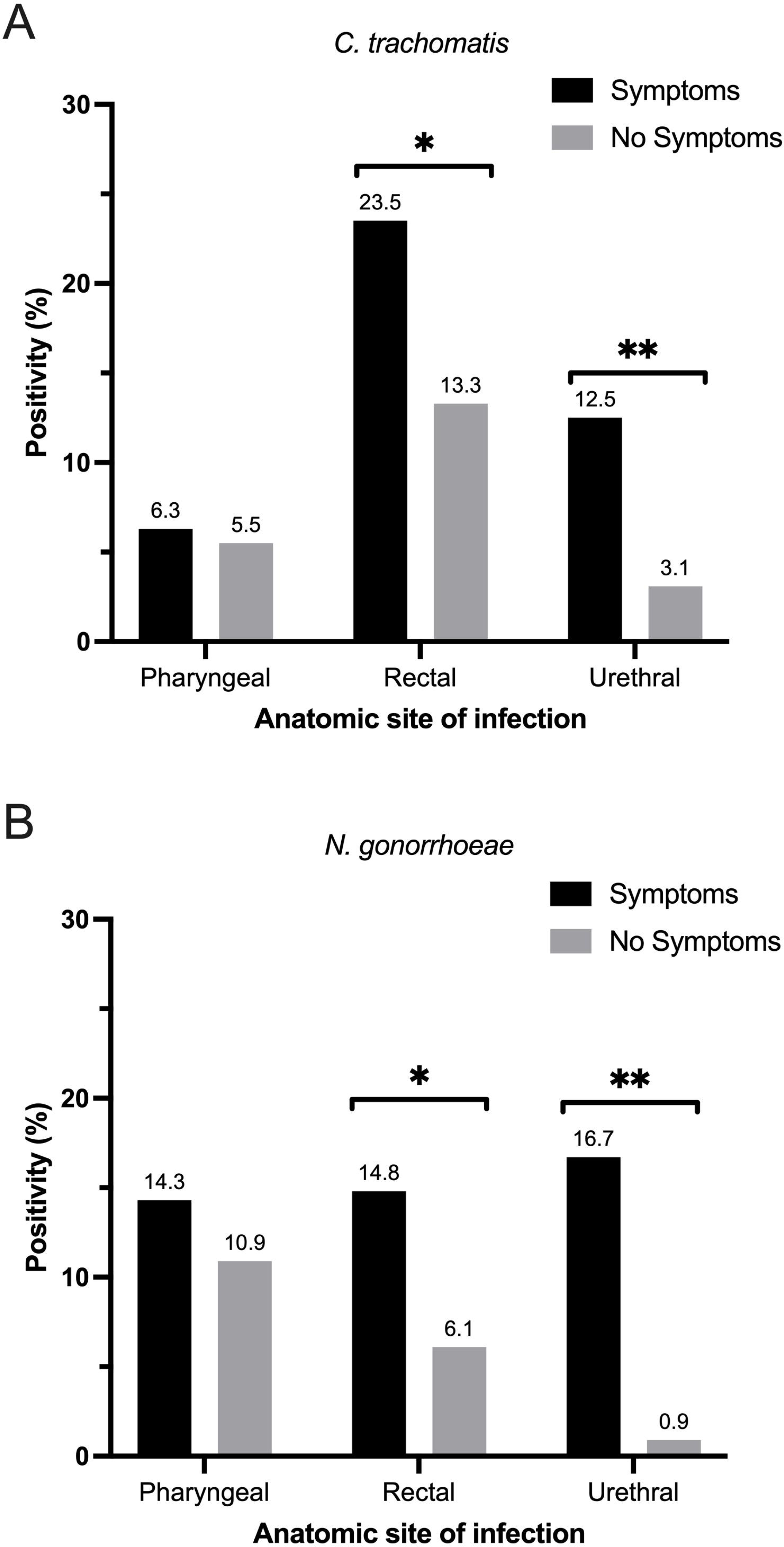
Prevalence of *C. trachomatis* (Panel A) and *N. gonorrhoeae* (Panel B) infections at pharyngeal, rectal, and urethral sites, stratified by reported symptoms at that site in the prior week. Footnote: *p-value ≤0.05; **p-value ≤0.01

### Factors associated with *C*. trachomatis and N. gonorrhoeae infections

In the multivariate logistic regression models, having either *C. trachomatis* or *N. gonorrhoeae* infection was associated with condomless anal intercourse in the prior month (aOR: 1.98; 95% CI: 1.27-3.09) and sexualized drug use in the prior 6 months (aOR: 1.71; 95% CI: 1.09, 2.69). (Table 3) Rectal and urethral symptoms in the prior week were associated with infections in univariate analyses, but not in the multivariate model (aOR: 1.19; 95% CI: 0.73, 1.94). When examining each infection separately, meeting sex partners in mobile apps in the prior 6 months was an independent risk factor for *N. gonorrhoeae* infections (aOR: 2.32; 95% CI: 1.19, 4.53), but not *C. trachomatis* (aOR: 0.83; 95% CI: 0.49, 1.43). Both condomless anal sex in the prior month (aOR: 2.10; 95% CI: 1.27-3.45) and sexualized drug use in the prior 6 months (aOR: 2.20; 95% CI: 1.32-3.68) were associated with *C. trachomatis*, but not *N. gonorrhoeae*. There was no evidence of multicollinearity among variables in the multivariate logistic regression.

**Table 3.** Multivariable logistic regression with factors associated with *C. trachomatis* only, *N. gonorrhoeae* only, and *C. trachomatis*or *N. gonorrhoeae* infections.

| Characteristics | *C. trachomatis* |  | *N. gonorrhoeae* |  | *C. trachomatis* or *N. gonorrhoeae* |  |
| --- | --- | --- | --- | --- | --- | --- |
|  | aOR (95% CI) | p-value | aOR (95% CI) | p-value | aOR (95% CI) | p-value |
| **Age group** |  |  |  |  |  |  |
| 16 to < 25 years old | 1.00 |  | 1.00 |  | 1.00 |  |
| 25 to < 35 years old | 0.75 (0.45, 1.27) | 0.284 | 1.03 (0.58, 1.84) | 0.912 | 0.81 (0.51, 1.30) | 0.387 |
| ≥ 35 years old | 1.27 (0.58, 2.80) | 0.550 | 0.39 (0.11, 1.38) | 0.144 | 0.76 (0.36, 1.64) | 0.492 |
| **Two or more sex partners in prior 1 month** | 1.66 (0.97, 2.86) | 0.066 | 1.16 (0.63, 2.14) | 0.638 | 1.52 (0.94, 2.46) | 0.086 |
| **Condomless anal sex in prior 1 month** | **2.10 (1.27, 3.45)** | **0.004** | 1.51 (0.86, 2.67) | 0.152 | **1.98 (1.27, 3.09)** | **0.002** |
| **Sexualized drug use in prior 6 months** | **2.20 (1.32, 3.68)** | **0.003** | 1.09 (0.61, 1.94) | 0.774 | **1.71 (1.09, 2.69)** | **0.020** |
| **Met sex partners via mobile apps** | 0.83 (0.49, 1.43) | 0.513 | **2.32 (1.19, 4.53)** | **0.013** | 1.21 (0.74, 1.96) | 0.445 |
| **Any anal or urethral symptoms in prior week** | 1.44 (0.84, 2.45) | 0.185 | 1.30 (0.72, 2.36) | 0.385 | 1.19 (0.73, 1.94) | 0.491 |
| **Used an antiseptic mouthwash in the prior 1 month** | 1.15 (0.70, 1.87) | 0.583 | 0.72 (0.41, 1.25) | 0.240 | 0.85 (0.55, 1.32) | 0.468 |
| **Model information** |  |  |  |  |  |  |
| Sample size included | 396 |  | 396 |  | 396 |  |
| Hosmer-Lemeshow p-value | 0.5244 |  | 0.5412 |  | 0.4685 |  |
| McFadden's R2 | 0.070 |  | 0.053 |  | 0.053 |  |
| Log likelihood | -202.98235 |  | -165.80362 |  | -237.93177 |  |

## Discussion

In this study including 529 primarily MSM in an HIV PrEP program in Hanoi, Vietnam, testing for *C. trachomatis* and *N. gonorrhoeae* at multiple anatomic sites identified a high prevalence (29%) of infections. Rectal infections were most common for *C. trachomatis* (14%), while pharyngeal infections were most common for *N. gonorrhoeae* (11%), demonstrating the utility of testing at multiple anatomic sites for the detection of infections outside of the genital tract. While the prevalence of urethral and rectal *C. trachomatis* and *N. gonorrhoeae* infections were higher among those who reported symptoms at those sites in the prior week, a majority of overall infections (54%) were detected among those without symptoms, underscoring the limitations of relying on syndromic management as an infection control strategy in this setting. These findings highlight the opportunity to enhance STI diagnosis and treatment within HIV PrEP programs in LMICs.

Our study provides important data on routine testing for *C. trachomatis* and *N. gonorrhoeae* within a large HIV PrEP program in a LMIC setting, where prevalence data are limited. A recent meta-analysis of STIs among people in HIV PrEP programs estimated a pooled prevalence of *C. trachomatis* or *N. gonorrhoeae* was 24%, suggesting a slightly higher burden among participants in our study population.[5] However, that study highlighted the scarcity of data from LMICs, which makes comparisons difficult, but the study did not find a significant difference in prevalence of chlamydia or gonorrhea by country income level. There is a paucity of published prevalence estimates of *C. trachomatis* and *N. gonorrhoeae* among MSM on HIV PrEP from other nearby countries in Asia, which could provide further context. One study done among adolescent MSM and transgender women in Thailand found a *C. trachomatis* prevalence of 15% and *N. gonorrhoeae* prevalence of 4.5%, estimates that are much lower than our study, although pharyngeal testing was not performed in that study which would likely account for some of that difference.[11] The prevalence of infections observed in our study align with existing evidence from other, largely high-income, settings indicating a high burden of STIs among MSM on HIV PrEP.[6,12]

Among participants in our study, we observed those reporting condomless anal sex were about twice as likely to have *C. trachomatis* or *N. gonorrhoeae* infection and those having sexualized drug use in the prior 6 months were 70% more likely to have *C. trachomatis* or *N. gonorrhoeae* infection. Identifying factors associated with infections could help to focus testing among those at highest risk for infections. For example, of the total 153 *C. trachomatis* or *N. gonorrhoeae* infections in our study, testing those reporting either condomless anal sex or sexualized drug use in the prior 6 months would have detected 75% of those infections. Meeting sex partners on mobile apps is increasingly common globally and this behavior was associated with twice the odds of *N. gonorrhoeae*, but not *C. trachomatis*, infections in our study, a finding that was also observed in our previous work in a similar study population.[8] Possible explanations for this observation might be there is a higher prevalence of *N. gonorrhoeae* among certain sexual networks of online mobile app users or there is more oral sex within that network, since the majority of these infections were pharyngeal. These findings indicate that focused testing for *N. gonorrhoeae* among those meeting sex partners on apps could be a strategy to improve gonorrhea control.

We found that reporting rectal or urethral symptoms in the prior week was not associated with infections in multivariate analysis, highlighting the difficulty of using symptoms to identify infections, given a high proportion of these types of infections are asymptomatic.[13] Despite this, the prevalence of rectal and urethral *C. trachomatis* and *N. gonorrhoeae* was significantly higher among those who reported symptoms in the prior week, suggesting symptoms are still correlated with a higher frequency of infections at those sites. Our findings suggest that testing for urethral infections among asymptomatic individuals is of limited value, as the prevalence in this group was very low, 0.9% for *N. gonorrhoeae* and 3.1% for *C. trachomatis*; furthermore, testing only at urethral sites would have missed more than three-quarters (77.8%) of these infections. The low positivity coupled with the high costs of testing, approximately 25-32 USD for one site *C. trachomatis/N. gonorrhoeae* testing in Vietnam, are important considerations, especially as the costs are typically managed by the patient, outside of research settings.

Pooled testing, where samples from one individual from multiple anatomic sites are combined into one sample for testing, is an approach that might be cost-saving but is associated with some compromise in test sensitivity.[14,15] However, anatomic sites of infection are important for treatment and follow-up, particularly with regard to antimicrobial resistance in *N. gonorrhoeae*, which would limit the cost savings of this approach in a high prevalence setting such as ours. Further research on the implementation and cost-effectiveness of pooled testing within HIV PrEP programs in LMICs would help to provide further guidance.

Our study further highlights the high prevalence of *C. trachomatis* and *N. gonorrhoeae* infections among people in HIV PrEP programs and the need to improve evidence-based guidelines for testing, particularly in LMICs. Given the high prevalence and incidence of STIs in HIV PrEP programs, many guidelines recommend quarterly screening for *C. trachomatis* and *N. gonorrhoeae* among MSM on HIV PrEP.[16,17] Yet while some modeling studies support quarterly screening to reduce the population prevalence, the clinical and public health evidence supporting the frequency of testing is lacking and the populations coverage needed to achieve a population-benefit of decreased prevalence is much higher than observed in practice.[18–22]

The recent Gonoscreen Study, a randomized-controlled trial evaluating quarterly screening for *C. trachomatis* and *N. gonorrhoeae* compared to no screening among MSM in Belgium found quarterly screening was associated with fewer *C. trachomatis* infections and complications, but observed no difference among *N. gonorrhoeae* infections.[23] Additional research on alternative screening frequencies, e.g. – every 6 months or annually, and in different settings could provide further evidence on the public health benefits of different screening approaches. Beyond the frequency of screening, there is also a larger debate about the utility of screening for asymptomatic STIs in populations of MSM, particularly those on HIV PrEP.[24,25] While *C. trachomatis* and *N. gonorrhoeae* infections increase the risk for HIV transmission and provide a strong rationale for using these as an entry point for HIV PrEP,[26] the increased risk for HIV acquisition caused by bacterial STIs is largely mitigated among those on HIV PrEP.[27,28] Moreover, antimicrobial resistance is one potential consequence of increased screening and treatment and is a significant concern, particularly for *N. gonorrhoeae* and *Mycoplasma genitalium*; increased antibiotic consumption was observed among those randomized to quarterly screening in the Gonoscreen study.[23] Antimicrobial resistance in *N. gonorrhoeae* is particularly concerning in Vietnam, where the prevalence of ceftriaxone resistance, the first-line agent for treatment, is more than 20% in some settings.[29]

Our study findings must be interpreted in light of the following limitations. First, the study population might have been subject to selection bias. Efforts were made to expand inclusivity by simplifying the study eligibility criteria to reflect that of the HIV PrEP program. Still, it was not possible to recruit all PrEP program clients and 70% of those who were recruited agreed to be screened; thus, it is possible those who chose not to participate were different than those who participated. Second, our study outcome was the prevalence of *C. trachomatis* and *N. gonorrhoeae* infections at each visit, so while participants were allowed to participate more than once, few did, and we were unable to estimate the incidence of *C. trachomatis* or *N. gonorrhoeae* infections. Third, our study population of primarily MSM were largely well-educated, employed, and living in Hanoi, the second most populated city in Vietnam, and might not be generalizable to other populations or settings. Our study took place within the largest HIV PrEP program in Hanoi, which is a strength of our study.

## Conclusion

Our findings provide further evidence of the high prevalence of *C. trachomatis* and *N. gonorrhoeae* infections among people on HIV PrEP, including the high proportions of rectal, pharyngeal, and asymptomatic infections. Despite the high prevalence of infections, the high cost of molecular STI testing is a major barrier to implementing routine testing in HIV PrEP programs in LMICs.[6] Testing for urethral *C. trachomatis* and *N. gonorrhoeae* infections among those without urethral symptoms is likely to be of limited value, based on our findings. Further research is needed to establish evidence-based recommendations on the role and frequency of testing for asymptomatic infections, including the cost-effectiveness of different approaches.

Biomedical interventions like doxycycline prophylaxis and vaccinations are promising tools for STI prevention and further research is needed on the effectiveness and implementation in LMIC settings, where key factors such as antimicrobial resistance and differences in user preferences will influence their impact.

## Data Availability

All data produced in the present study are available upon reasonable request to the authors.

## Acknowledgements

The research was supported by a grant of no charge materials from Roche Molecular Systems and Abbott Molecular. Authorship: PCA, HTB, GML, and JDK designed the study. PCA, GML, JDK obtained funding. HTB and LQP performed data management and data analyses. PCA wrote the first version of the manuscript. All authors reviewed, provided critical review, and approved the manuscript.

## References

1 World Health Organization. Global progress report on HIV, viral hepatitis and sexually transmitted infections, 2021. Geneva, Switzerland.: World Health Organization; 2021. https://www.who.int/publications/i/item/9789240027077 (accessed 20 Jul2024).

2 Centers for Disease Control and Prevention. Sexually Transmitted Infections Surveillance 2022. Atlanta: US Department of Health and Human Services; 2024. https://www.cdc.gov/std/statistics/2022/default.htm (accessed 12 May2024).

3 Kojima N, Davey DJ, Klausner JD. Pre-exposure prophylaxis for HIV infection and new sexually transmitted infections among men who have sex with men. AIDS 2016; 30:2251– 2252.

4 Traeger MW, Schroeder SE, Wright EJ, Hellard ME, Cornelisse VJ, Doyle JS, et al. Effects of Pre-exposure Prophylaxis for the Prevention of Human Immunodeficiency Virus Infection on Sexual Risk Behavior in Men Who Have Sex With Men: A Systematic Review and Meta-analysis. Clin Infect Dis 2018; 67:676–686.

5 Ong JJ, Baggaley RC, Wi TE, Tucker JD, Fu H, Smith MK, et al. Global Epidemiologic Characteristics of Sexually Transmitted Infections Among Individuals Using Preexposure Prophylaxis for the Prevention of HIV Infection: A Systematic Review and Meta-analysis. JAMA Netw Open 2019; 2:e1917134.

6 Ong JJ, Fu H, Baggaley RC, Wi TE, Tucker JD, Smith MK, et al. Missed opportunities for sexually transmitted infections testing for HIV pre-exposure prophylaxis users: a systematic review. J Int AIDS Soc 2021; 24:e25673.

7 Wi TE, Ndowa FJ, Ferreyra C, Kelly-Cirino C, Taylor MM, Toskin I, et al. Diagnosing sexually transmitted infections in resource-constrained settings: challenges and ways forward. J Int AIDS Soc 2019; 22 Suppl 6:e25343.

8 Adamson PC, Bhatia R, Tran KDC, Bui HTM, Vu D, Shiraishi RW, et al. Prevalence, Anatomic Distribution, and Correlates of Chlamydia trachomatis and Neisseria gonorrhoeae Infections Among a Cohort of Men Who Have Sex With Men in Hanoi, Vietnam. Sex Transm Dis 2022; 49:504–510.

9 Vietnam Authority for HIV Prevention and Control. Guideline on implementation HIV preexposure prophylaxis (PrEP) services (Document No. 133). Vietnam Ministry of Health.; 2020.

10 Nevendorff L, Schroeder SE, Pedrana A, Bourne A, Stoové M. Prevalence of sexualized drug use and risk of HIV among sexually active MSM in East and South Asian countries: systematic review and meta-analysis. Journal of the International AIDS Society 2023; 26:e26054.

11 Songtaweesin WN, Pornpaisalsakul K, Kawichai S, Wacharachaisurapol N, Wongharn P, Yodkitudomying C, et al. Sexually transmitted infections incidence in young Thai men who have sex with men and transgender women using HIV pre-exposure prophylaxis. Int J STD AIDS 2022; 33:447–455.

12 World Health Organization. WHO implementation tool for pre-exposure prophylaxis (PrEP) of HIV infection. Module 13. Integrating STI services. Geneva, Switzerland.:; 2022. https://www.who.int/publications/i/item/9789240057425 (accessed 11 Jun2024).

13 Kent CK, Chaw JK, Wong W, Liska S, Gibson S, Hubbard G, et al. Prevalence of Rectal, Urethral, and Pharyngeal Chlamydia and Gonorrhea Detected in 2 Clinical Settings among Men Who Have Sex with Men: San Francisco, California, 2003. Clinical Infectious Diseases 2005; 41:67–74.

14 Aboud L, Xu Y, Chow EPF, Wi T, Baggaley R, Mello MB, et al. Diagnostic accuracy of pooling urine, anorectal, and oropharyngeal specimens for the detection of Chlamydia trachomatis and Neisseria gonorrhoeae: a systematic review and meta-analysis. BMC Med 2021; 19:285.

15 Almeria J, Pham J, Paris KS, Heskett KM, Romyco I, Bristow CC. Pooled 3-Anatomic-Site Testing for Chlamydia trachomatis and Neisseria gonorrhoeae: A Systematic Review and Meta-Analysis. Sexually Transmitted Diseases 2021; 48:e215.

16 Centers for Disease Control and Prevention. Preexposure prophylaxis for the prevention of HIV infection in the United States—2021 Update: a clinical practice guideline.; 2021. https://www.cdc.gov/hiv/pdf/risk/prep/cdc-hiv-prep-guidelines-2021.pdf (accessed 12 Jun2024).

17 British HIV Association, British Association of Sexual Health and HIV. BHIVA/BASHH guidelines on the use of HIV pre-exposure prophylaxis (PrEP), 2018.; 2018. https://www.bhiva.org/PrEP-guidelines (accessed 12 Jun2024).

18 Jenness SM, Weiss KM, Goodreau SM, Gift T, Chesson H, Hoover KW, et al. Incidence of Gonorrhea and Chlamydia Following Human Immunodeficiency Virus Preexposure Prophylaxis Among Men Who Have Sex With Men: A Modeling Study. Clinical Infectious Diseases 2017; 65:712–718.

19 Reitsema M, van Hoek AJ, van der Loeff MS, Hoornenborg E, van Sighem A, Wallinga J, et al. Preexposure prophylaxis for men who have sex with men in the Netherlands: impact on HIV and Neisseria gonorrhoeae transmission and cost-effectiveness. AIDS 2020; 34:621.

20 Hocking JS, Temple-Smith M, Guy R, Donovan B, Braat S, Law M, et al. Population effectiveness of opportunistic chlamydia testing in primary care in Australia: a cluster-randomised controlled trial. The Lancet 2018; 392:1413–1422.

21 Tsoumanis A, Hens N, Kenyon CR. Is Screening for Chlamydia and Gonorrhea in Men Who Have Sex With Men Associated With Reduction of the Prevalence of these Infections? A Systematic Review of Observational Studies. Sexually Transmitted Diseases 2018; 45:615.

22 Tao G, Patel CG, He L, Workowski K. STI/HIV testing, STIs, and HIV PrEP use among men who have sex with men (MSM) and men who have sex with men and women (MSMW) in United States, 2019-2022. Clin Infect Dis 2024;: ciae314.

23 Vanbaelen T, Tsoumanis A, Florence E, Van Dijck C, Huis in’t Veld D, Sauvage A-S, et al. Effect of screening for Neisseria gonorrhoeae and Chlamydia trachomatis on incidence of these infections in men who have sex with men and transgender women taking HIV pre-exposure prophylaxis (the Gonoscreen study): results from a randomised, multicentre, controlled trial. The Lancet HIV 2024; 11:e233–e244.

24 Williams E, Williamson DA, Hocking JS. Frequent screening for asymptomatic chlamydia and gonorrhoea infections in men who have sex with men: time to re-evaluate?The Lancet Infectious Diseases 2023; 23:e558–e566.

25 Ridpath AD, Chesson H, Marcus JL, Kirkcaldy RD, Torrone EA, Aral SO, et al. Screening Peter to Save Paul: The Population-Level Effects of Screening Men Who Have Sex With Men for Gonorrhea and Chlamydia. Sexually Transmitted Diseases 2018; 45:623.

26 Kasaie P, Schumacher CM, Jennings JM, Berry SA, Tuddenham SA, Shah MS, et al. Gonorrhoea and chlamydia diagnosis as an entry point for HIV pre-exposure prophylaxis: a modelling study. BMJ Open 2019; 9:e023453.

27 Volk JE, Marcus JL, Phengrasamy T, Blechinger D, Nguyen DP, Follansbee S, et al. No New HIV Infections With Increasing Use of HIV Preexposure Prophylaxis in a Clinical Practice Setting. Clin Infect Dis 2015; 61:1601–1603.

28 Hoornenborg E, Coyer L, Achterbergh RCA, Matser A, Schim van der Loeff MF, Boyd A, et al. Sexual behaviour and incidence of HIV and sexually transmitted infections among men who have sex with men using daily and event-driven pre-exposure prophylaxis in AMPrEP: 2 year results from a demonstration study. The Lancet HIV 2019; 6:e447–e455.

29 Adamson PC, Hieu VN, Nhung PH, Whiley DM, Chau TM. Ceftriaxone resistance in Neisseria gonorrhoeae associated with the penA-60.001 allele in Hanoi, Viet Nam. The Lancet Infectious Diseases 2024; 0. doi:10.1016/S1473-3099(24)00230-5

